# Hypomethylation on cfDNA within transposable elements can predict ovarian malignancy in women with adnexal masses

**DOI:** 10.64898/2026.01.14.26344128

**Authors:** Melanie Weigert, Xiao-Long Cui, Peihong Zhang, Krissana Kowitwanich, Diana West-Szymanski, Sarah Rauch, Wei Zhang, Chuan He, Ernst Lengyel

## Abstract

**Objective:** Tumor transformation is accompanied by widespread methylation changes, in the form of cytosine modifications, in transposable elements (TEs). The purpose of this study was to investigate whether changes in methylation and hydroxymethylation in the form of 5mC and 5hmC within transposable elements in circulating cell-free DNA (cfDNA) can serve as predictive markers of ovarian malignancy.

**Methods:** Healthy women as well as women undergoing surgery for various benign, borderline, or malignant adnexal masses and gynecologic conditions were selected for this study. 5hmC-Seal or LABS-seq were performed on cfDNA isolated from prospectively collected serum and plasma samples. We built two models using either differentially hydroxymethylated (5hmC) or methylated (5mC) features within TEs to establish models predictive of ovarian malignancy in women with adnexal masses.

**Results:** We isolated cfDNA and analyzed a total of 522 samples (plasma, n = 363; serum, n = 159) from women receiving care at the University of Chicago Medical Center. Multivariate modelling using age, cell deconvolution and the top 38 differentially hydroxymethylated (5hmC) transposable elements was able to accurately predict malignancy in women with an area under the curve (AUC) of 0.854 and a 95% confidence interval: 0.746- 0.962 (95% CI) in plasma and an AUC of 0.893 (95% CI: 0.806- 0.98%) in serum. 5mC-based modelling using the top 25 positive and negatively correlated features, based on differentially methylated transposable element promoters, was able to accurately predict malignancy with an AUC of 0.970 (accuracy of 0.936).

**Conclusions:** We developed two multivariate models based on cfDNA-derived 5hmC or 5mC modified transposable elements to accurately predict ovarian malignancy in women. Furthermore, we show that our 5hmC-based model applies to plasma (EDTA) and serum samples, which are commonly collected in clinical practice. The results of our study warrant further investigation of the predictive performance of our models in large-scale cohorts in the future.

## Introduction

The frequent overuse of imaging modalities in medical practice has led to an increase in the detection of adnexal masses. Although adnexal masses are found in 35% of premenopausal and 17% of postmenopausal women (1), they pose a significant diagnostic challenge for gynecologists. Annually, over 200,000 women in the United States undergo surgical evaluation for an adnexal mass. While 90% of adnexal masses are benign, the risk of malignancy must be evaluated each time due to the high mortality associated with ovarian cancer (OvCa) (2,3), which necessitates early surgical intervention for long-term survival. Nevertheless, because the incidence of OvCa is low, many surgeries for an adnexal mass can be viewed as unnecessary and expose patients to the risk of surgical complications (4). Current clinical assays that utilize blood-based biomarkers to help evaluate adnexal masses include CA-125 testing alone, in combination with clinical parameters (e.g., menopause status) (5–8), and the second-generation multivariate index assay (MIA2G or OVERA), which measures multiple protein markers (9–11). However, all current tests vary widely in their sensitivity (78%-93%) and consistently fail to achieve high specificity (8,12–14). An accurate biomarker that can reliably assess the preoperative risk of ovarian malignancy would lead to more effective patient stratification for further diagnostic workups.

Extensive research over the past years has shown that malignant transformation is accompanied by widespread epigenetic changes, such as DNA methylation, which play pivotal roles in the onset and progression of malignancy (15–17). Previous studies have demonstrated that methylation patterns, in the form of 5-methylcytosine (5mC) and 5-hydroxymethylcytosine (5hmC), are stable and conserved in cell-free DNA (cfDNA). Several technologies, such as LABS-seq and nano-5hmC-Seal, that can interrogate methylation changes in cell-free DNA have revealed that aberrant methylation patterns within gene promoters (e.g., tumor suppressors) are promising biomarkers for cancer detection, diagnosis, and prognosis (18–21). Interestingly, most of the methylation found within the genome is concentrated within transposable elements (TE), such as retrotransposons, where methylation suppresses their transcriptional activity, which can otherwise result in genomic instability and chromosomal rearrangements (22). Studies have shown that the loss of methylation within the promoter regions of these elements is a widespread hallmark of several cancers (23–25). Because TEs are heavily methylated and highly abundant within the genome, we sought to assess whether methylation changes on cfDNA within TEs can provide highly sensitive biomarkers that can predict ovarian malignancy in women diagnosed with an adnexal mass.

We report that genome-wide profiling of 5hmC and 5mC on cfDNA in TEs can predict ovarian malignancy with high accuracy and provide novel minimally invasive diagnostic biomarkers.

## Materials and Methods

### Study population and inclusion criteria

Women aged 18 to 89 with no prior cancer history who underwent surgery for ovarian cancer or benign gynecologic conditions at The University of Chicago Medical Center were included in this study. The study was approved by the University of Chicago Institutional Review Board (Protocol# 13-372B and 21-1114), and informed consent was obtained from all study participants before participation.

### Cohort characteristics and study definitions

All study participants had their clinico-pathological data prospectively collected and stored in a REDCap database. Patient metadata are described in **Table S1**.

All patients were categorized into one of four categories: healthy, benign, borderline, and malignant based on their final surgical pathology. The malignant ovarian epithelial group includes serous (grade 1-3), mucinous, endometrioid, clear cell, synchronous, and carcinosarcomas of ovarian or fallopian tube origin. The malignant ovarian non-epithelial group includes germ cell tumors, while the malignant non-GYN metastasis group encompasses gastrointestinal (e.g., appendiceal) cancers that metastasized to the ovary. The malignant GYN cancer groups include endometrial cancers (e.g., serous or endometrioid) with or without ovarian involvement. The benign ovarian epithelial group includes Brenner tumors, as well as mucinous and serous cysts, while the benign ovarian non-epithelial group consists of teratomas, fibromas, and both follicular and hemorrhagic corpus luteum cysts. The benign inflammation group includes hydrosalpinx, pelvic inflammatory disease, and florid xanthogranulomatous inflammatory processes. The healthy group comprises germline mutation carriers (e.g., *BRCA1/2*, *PALB2, MSH2*, *RAD51C*, *BRIP1*) who underwent risk-reducing surgeries, as well as patients who underwent surgeries for uterine prolapse or abdominal cerclages. The benign extra-ovarian or uterine group includes fibroids and paratubal cysts. The borderline group consists of patients with adnexal masses of low malignant potential.

### Blood collection, processing, and cfDNA isolation

All blood samples were collected prospectively before surgery. Serum was drawn into red top tubes, left to clot for 30 min at room temperature, and then centrifuged at 3,000 rpm at 4°C for 11 min to separate the serum from the clot. Plasma was collected in purple top EDTA tubes and processed using a two-step centrifugation protocol. First, the blood was spun at 2,500 rpm at 4°C for 15 minutes to isolate the plasma. The plasma was carefully transferred to a clean tube without disturbing the buffy coat, then spun for a second time at 11,800 rpm at 4°C for 15 min. Samples were transferred to clean cryotubes and stored at −80°C. Cell-free DNA was extracted from serum or plasma using the QIAamp Circulating Nucleic Acid kit (Qiagen), following the manufacturer’s instructions, and stored at −20°C for short-term storage.

### Preparation and sequencing of nano-5hmC-Seal libraries

Six to ten ng of cfDNA were used to construct nano-5hmC-Seal libraries as previously described (21,26). In summary, 5hmC modifications on cfDNA were labeled enzymatically with UDP-N_3_-glucose (Active Motif) using T4 β-glucosyltransferase (Thermo Fisher) and chemically modified with biotin-PEG4-dibenzocyclooctyne (DBCO, Vector Laboratories). Biotin-labeled, 5hmC-specific pull-down was performed using streptavidin M270 Dynabeads (Invitrogen). The resulting libraries were PCR amplified before sequencing on an Illumina NovaSeq 6000 using fifty base pairs, paired-end sequencing. Sequence quality was assessed using FASTQC (27), version 0.11.9, before raw read trimming using Trim Galore (RRID: SCR_011847) and mapping to the human genome (hg19) using Bowtie2 (RRID: SCR_016368), version 2.2.5 (28). Following the removal of PCR duplicates using Samtools (RRID: SCR_002105), version 1.14 (29), uniquely mapped reads were used for downstream analysis.

### 5hmC distribution by genomic features

FeatureCounts version 2.0.1 was used to count reads across different genomic features. Enhancer regions were selected from the GeneHancer database (30) (RRID: SCR_023953), and CpG islands were extracted from the UCSC Genome Browser (University of California, Santa Cruz) (31). DESeq2-normalized (RRID: SCR_000154) read counts on the different genomic features were used for comparison, and differences in 5hmC levels between groups and by genomic features were assessed using the Kruskal-Wallis test.

### Functional enrichment analysis of genes with differential levels of 5hmC modifications

Reads aligned on gene bodies (mapping score ≥ 10) were calculated using FeatureCounts (32), followed by read count comparison using DESeq2, version 1.34.0 (33). The Pheatmap R package, version 1.0.12, was used to calculate distance matrices for hierarchical clustering and heatmap drawing. Differentially hydroxymethylated genes with a p-value < 0.05 were used to perform functional enrichment analysis in Metascape (34).

### Analysis of transposable elements

Genomic location information of different repeat elements was extracted from the RepeatMasker table of the UCSC Genome Browser. Reads aligned to the hg19 reference genome were counted on each repeat element using featureCounts, allowing multi-mapped reads. The DESeq2 R package was then used to normalize the read counts and find the differentially hydroxymethylated repeat elements.

### Development of a 5hmC model for malignancy prediction

All benign and malignant plasma samples were used to build and train the model. The samples were split into a training set (70%) and a validation set (30%). Normalized read counts were utilized to calculate the coefficient of variation for both benign and malignant samples. After batch correction, the top 50,000 age, BMI, and ethnicity-corrected transposable elements with the highest coefficients of variation were selected for feature selection. Elastic net with 5-fold cross-validation, optimized for the area under the curve, was performed 100 times to construct a linear regression model. After calculating feature frequencies, the top 10 transposable elements were selected for the final model (**Table S3**). The final model was applied to serum-derived benign and malignant samples for classification.

### Linear amplification-based bisulfite sequencing library preparation and data processing

LABS-seq libraries were prepared as previously described (20). In brief, cfDNA was ligated to T7 adaptors using the KAPA Hyper prep kit (Roche); the adapter-ligated cfDNA was cleaned using AMPure XP beads (Beckman Coulter), followed by bisulfite treatment and T7 primer extension. The resulting T7-tagged double-stranded DNA (dsDNA) fragments were then used as templates for *in vitro* transcription using the HiScribe T7 High Yield RNA Synthesis kit (NEB). To remove DNA templates, the reaction was digested using DNase I (NEB), and the remaining RNA templates were purified. 100 ng RNA template was used for library construction using the KAPA RNA HyperPrep kit (Roche) following the manufacturer’s instructions.

Raw reads were trimmed using Trim Galore (RRID: SCR_011847) and aligned to the human genome (hg19) using Bismark (35). The Methylit R package was used to determine the methylation proportion at each CpG and differentially methylated regions with a cutoff FDR < 0.01 and a difference of ≥15% were detected.

### 5mC distribution by genomic features

Palindromic cytosine-phosphate-linked-guanine (CpG) sites were annotated using *annotatePeaks.pl* tool from the HOMER package (v5.1) (36). Annotations were performed based on the UCSC genome assemblies, which HOMER uses internally to define gene features, CpG islands, and repetitive elements.

### Development of a 5mC model for malignancy prediction

To evaluate the predictive potential of 5mC within transposable elements, we constructed an XGBoost classifier and applied nested cross-validation to ensure robust and unbiased performance estimates (37,38). All samples were randomly divided into five stratified subsets with balanced numbers of benign and malignant patients. In each of five outer loops, one subset served as an independent test set, while the remaining samples underwent three-fold cross-validation for model training and hyperparameter tuning. Differentially methylated transposable promoters were used as input features.

Variation in feature importance across models suggested redundancy among differentially methylated promoters. To address this, we applied feature selection based on correlation with labels: benign and malignant samples were labeled 0 and 1, and Pearson correlation was computed between promoter methylation levels and labels. The top 25 positively and 25 negatively correlated promoters were selected for model training, yielding improved classification performance.

### Cell deconvolution analysis

CIBERSORT was used for the cellular deconvolution of genome-wide 5hmC and 5mC profiles. To construct a comprehensive 5hmC reference for cell deconvolution, 5hmC-Seal profiles from previously published normal tissues and cell types were analyzed (39,40). The top 100 genes for each tissue and cell type were selected and RPKM values calculated for both reference and cfDNA profiles, before deconvolution.

To construct a comprehensive 5mC reference, we used the whole genome bisulfite converted profiles of 40 different cell types (41). Methylation values for cell type-specific differentially methylated regions were used to deconvolute our cfDNA methylomes.

### Statistical analysis

All statistical analysis were performed in R project. Two-tailed tests were used for all analyses, and p< 0.05 and q< 0.05 were considered statistically significant.

### Data availability

Requests for resources, reagents, and additional information should be directed and will be fulfilled by the lead contact, Ernst Lengyel. De-identified human 5hmC and 5mC DNA sequencing data will be made available upon publication.

### Code availability

All software and packages used for analysis have been published previously. Any code used in this study can be shared upon request.

## Results

### Clinico-pathologic characteristics

The 5-hydroxymethylcytosine (5hmC) cohort included a total of 376 prospectively collected plasma (n = 217) and serum (n = 159) samples collected from women undergoing treatment at the University of Chicago Medical Center (**Figure 1**). The cohort consisted of 57 (15.2%) healthy women, 144 (38.3%) women who had surgery for benign gynecologic conditions, 32 women who underwent surgery for borderline ovarian tumors (8.5%), and 143 women who underwent surgery for malignant (38%) tumors. The mean ages were 50 years for healthy women, 50.9 years for women with benign disease, 53.5 years for women with borderline disease, and 60.4 years for women with malignant disease. Two hundred twenty-four women were of European descent (59.6%), 97 were African American (25.8%), 22 were Hispanic (5.9%), 11 were Asian (2.9%), and 22 women chose not to provide information (5.8%) in the validation cohort (**Table S1**).

**Figure 1:**
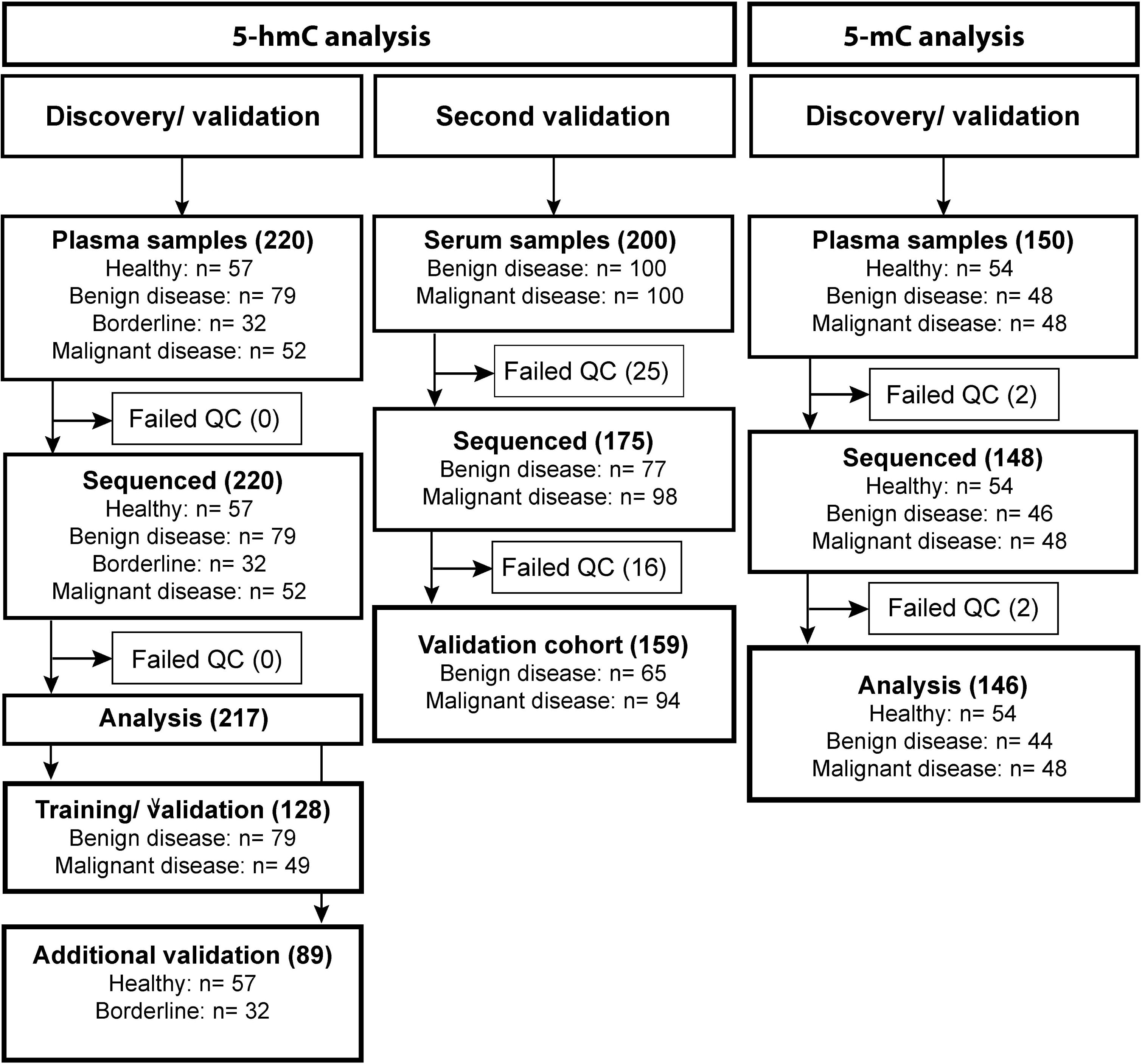
Overview of the study cohort and sample workflow. A total of 522 samples were analyzed in this study, including 159 serum and 363 plasma samples. After cfDNA isolation and library preparation, the 5hmC discovery/validation cohort included 79 benign and 49 malignant samples for training and validation, along with 57 healthy and 32 borderline plasma samples for further validation. The second 5hmC validation cohort consisted of 65 benign and 94 malignant serum samples. The 5mC discovery/validation cohort comprised 54 healthy, 44 benign, and 48 malignant plasma samples.

The 5-methylcytosine (5mC) cohort included a total of 146 plasma samples, of which 54 women were healthy (36.5%), 44 were diagnosed with benign disease (30.1%), and 48 had an invasive (33.4%) OvCa. The mean age for healthy women was 41.9 years, 41.5 years for women with benign disease, and 60.8 years for women with malignant disease. Seventy-nine women were of European descent (54.1%), 49 were African American (33.6%), 8 were Hispanic (5.5%), 4 belonged to other ethnicities (2.7%), and 6 declined (4.1%) to provide information (**Table S1**).

### 5hmC signals in plasma-derived cfDNA can differentiate benign and malignant disease

Cell-free DNA (cfDNA) was extracted from 220 plasma samples, followed by nano-hmC-Seal library preparation, sequencing, and data analysis, resulting in a plasma cohort consisting of 217 samples (**Table S1**). First, the distribution of 5hmC by genomic features was assessed and found that the majority of 5hmC reads were located within introns (32.89%) and intergenic regions (20%), followed by transposable elements (TE) such as LINE (10.7%) and SINE (7.69%) (**Figure 2A** and **Table S2**). Distribution enrichment by genomic features in all samples revealed an enrichment of 5hmC within exons, promoters, and transcription start sites, while a loss of 5hmC was observed within TEs like SINE and LINE (**Figure S1A**). Previous work has shown that 5hmC enrichment within gene bodies (i.e., promoters, TSS) can reveal insights into the biology and pathways underlying certain diseases (21,26,42). Hierarchical clustering revealed that differentially hydroxymethylated genes (DhMGs) could not distinguish between patients with benign and those with malignant gynecologic disease (**Figure S1B**). Next, functional enrichment analysis was performed to identify pathways that might drive and distinguish the two disease groups. Samples from patients with benign gynecologic conditions showed 5hmC enrichment in pathways associated with regulation of immune responses and responses to interleukin-4, while samples from patients with malignant disease showed enrichment in pathways involved in cell junction organization and trans-synaptic signaling (**Figure 2B**).

**Figure 2:**
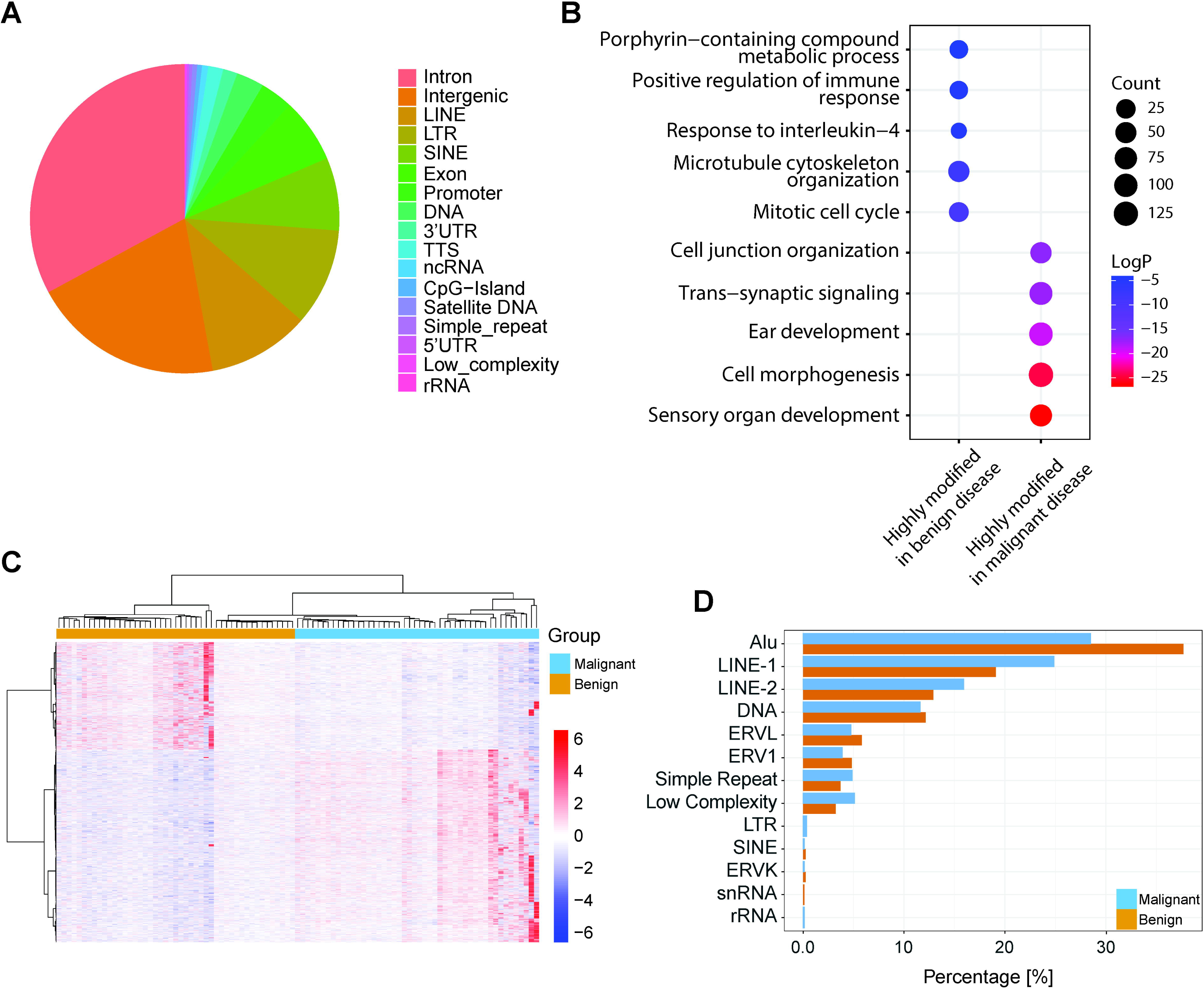
5hmC signals on plasma-derived cfDNA can distinguish between benign and malignant disease. A) 5hmC distribution across genomic features in benign (n=79) and malignant (n=49) samples. B) Functional enrichment analysis of benign and malignant samples based on all significantly differentially hydroxymethylated genes (DhmGs). C) Hierarchical clustering of 2,501 differentially hydroxymethylated transposable elements by disease. D) Distribution enrichment of 5hmC by transposable elements. LINE: long-interspersed nuclear element, SINE: short-interspersed nuclear element, LTR: long terminal repeat, TTS: transcription termination site, 3’UTR: 3’ prime untranslated region, CpG-Island: palindromic cytosine-phosphate-linked-guanine islands, ncRNA: non-coding RNA, and 5’UTR: 5’ prime untranslated region, Alu: Arthrobacter luteus, ERVL: endogenous retrovirus-like element, ERV1: endogenous retroviral sequence 1, ERVK: endogenous retrovirus type K, snRNA: small nuclear RNA and rRNA: ribosomal RNA.

Loss of methylation and reactivation of TEs are recognized hallmarks of certain cancers (24,43,44). Since we observed that 32.2% of our 5hmC reads were in TEs, we aimed to determine whether differential hydroxymethylation within these elements could be used to distinguish benign from malignant samples. Hierarchical clustering and principal component analysis (PCA) revealed that differentially hydroxymethylated TEs effectively separate benign from malignant samples (**Figure 2C** and **S1C-F**). Further enrichment analysis indicated that the majority of 5hmC reads in the benign group were derived from Alu elements (43.5%), whereas the majority of 5hmC reads in the malignant group originated from LINE-1 (20.67%) and LINE-2 (10.83%) elements (**Figure 2D** and **Table S3**). Cell deconvolution analysis showed that 5hmC signals from healthy and benign cases in plasma were significantly enriched in B cells compared with those from malignant cases (**Figure S2**).

### 5hmC within transposable elements can predict malignancy

Currently, there are no highly accurate tests or biomarkers that can predict ovarian malignancy. To address this gap, we aimed to develop and evaluate a model that predicts malignancy based on 5hmC modifications within TEs. Using linear regression, we built a model using age, cell deconvolution (B and CD8T cells) as well as the top 38 differentially hydroxymethylated TEs for prediction (**Figure 3A** and **Table S4**). The model achieved an AUC of 0.854 (95% CI, 0.746-0.962) for predicting malignancy in plasma samples (**Figure 3B**). Although our model was trained on benign and malignant samples, our plasma cohort also included an additional 57 healthy and 32 borderline cases. Using PCA, we examined whether healthy samples resemble benign samples and whether borderline samples are more like malignant samples. Indeed, healthy samples are more similar to benign ones, and borderline samples are more similar to malignant samples (**Figure 3C**). Previous analysis showed that serum- and plasma-derived 5hmC annotations were highly correlated and showed no significant differences, therefore final validation was performed using a second cohort consisting of serum samples. Cell deconvolution analysis of the serum samples revealed that like plasma samples (**Figure S2**), benign cases were enriched in 5hmC signals derived from B cells (**Figure S3**). The serum cohort included 65 benign and 94 malignant samples and achieved an AUC of 0.893 (95% CI, 0.806- 0.980) (**Figure 3D**). In summary, these data establish that 5hmC modifications within TEs can predict malignancy using both serum and plasma as an analyte.

**Figure 3:**
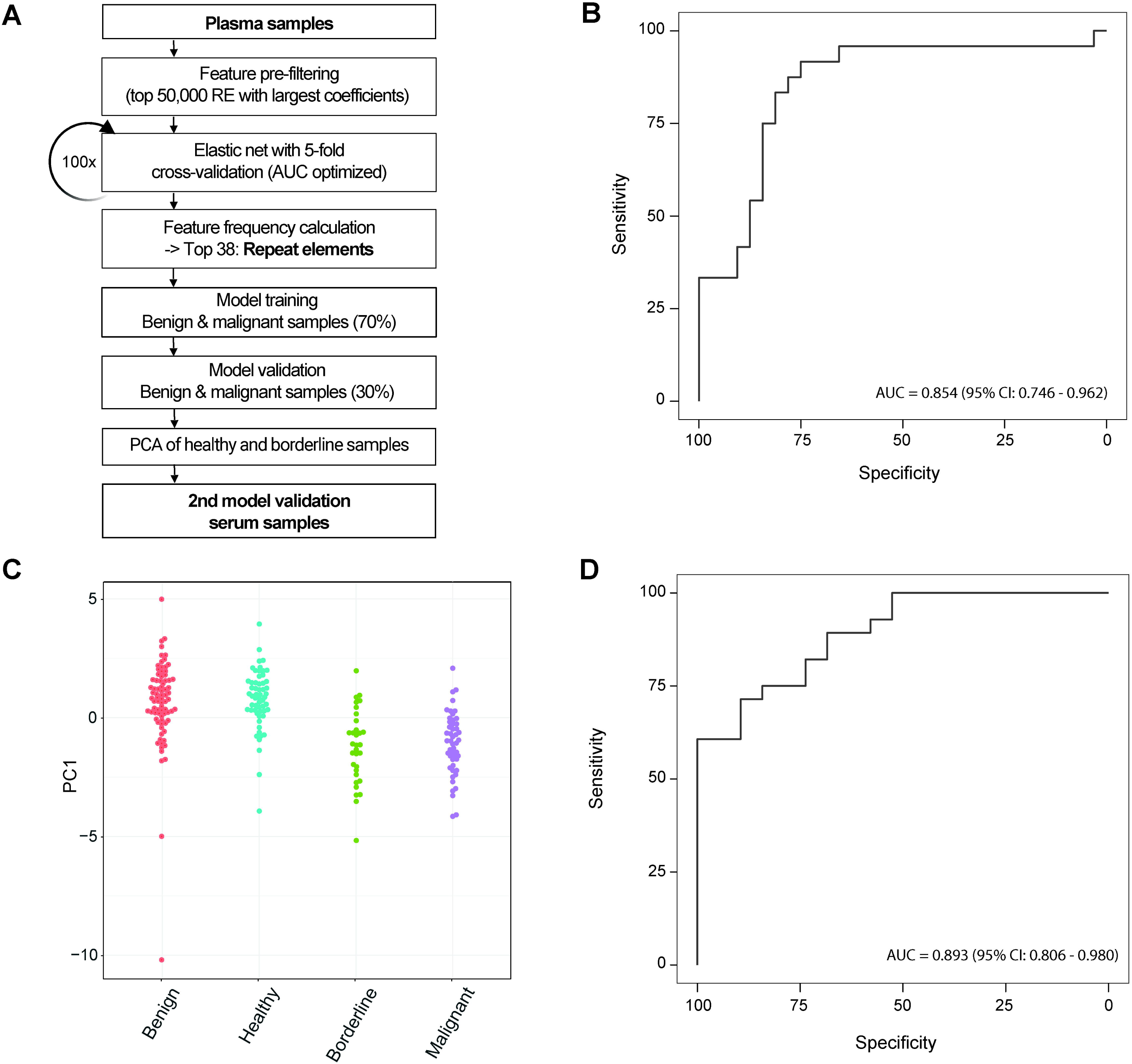
5hmC within transposable elements is a biomarker predictive of malignancy. A) Steps involved in building a model that can separate patients with benign (n= 79) from malignant (n= 49) disease based on plasma (training and validation set) and 159 serum (second validation set) samples. B) Receiver operating curves (ROC) optimized for the area under the curve (AUC) based on the top 38 differentially hydroxymethylated transposable elements that differentiate between benign and malignant disease. C) Principal component analysis (PCA) based on the top 38 transposable elements used to build the model shows that healthy samples (n= 57) are similar to benign samples, while borderline (n= 32) samples are similar to malignant samples. D) ROC based on serum samples in the second validation set containing patients with benign (n= 65) and malignant (n= 94) disease.

### Cell-free DNA methylation reveals global changes and copy number alterations

Tumorigenesis is associated with aberrant changes in 5mC methylation and loss thereof (45). Therefore, we performed linear amplification bisulfite sequencing (LABS-seq) (20) to assess 5mC levels on cfDNA extracted from healthy patients (n = 54), and patients with benign (n = 48) and malignant (n = 48) disease (**Figure 1**). Cell deconvolution analysis using 5mC revealed that malignant/ borderline cases were enriched in T cells and while not significant B cells showed a similar pattern of enrichment in healthy and benign cases compared to malignant/ borderline samples (**Figure S4**). Principal component analysis (PCA) of globally differentially methylated regions (DMRs) indicated a sequential transition from healthy to benign and eventually towards malignant cases (**Figure 4A**). Further analysis using hierarchical clustering based on 211 DMRs further revealed sequential loss of 5mC, indicating that global demethylation accompanies ovarian tumorigenesis (**Figure 4B**). This is consistent with prior research showing that tumorigenesis is often associated with aberrant methylation changes and its widespread loss (15,44). Copy number alterations (CNA) are a hallmark of OvCa (47). LABS-seq-based analysis of genome-wide 5mC modifications on cfDNA uncovered distinct CNAs (**Figure 4C**) along with the lengths of the genomic regions affected by CNA (**Figure 4D**). One patient within the benign group had a significant CNA (marked with an asterisk). Upon further review, we noted that this patient was later diagnosed with cervical intraepithelial neoplasia (CIN) after the initial blood draw.

**Figure 4:**
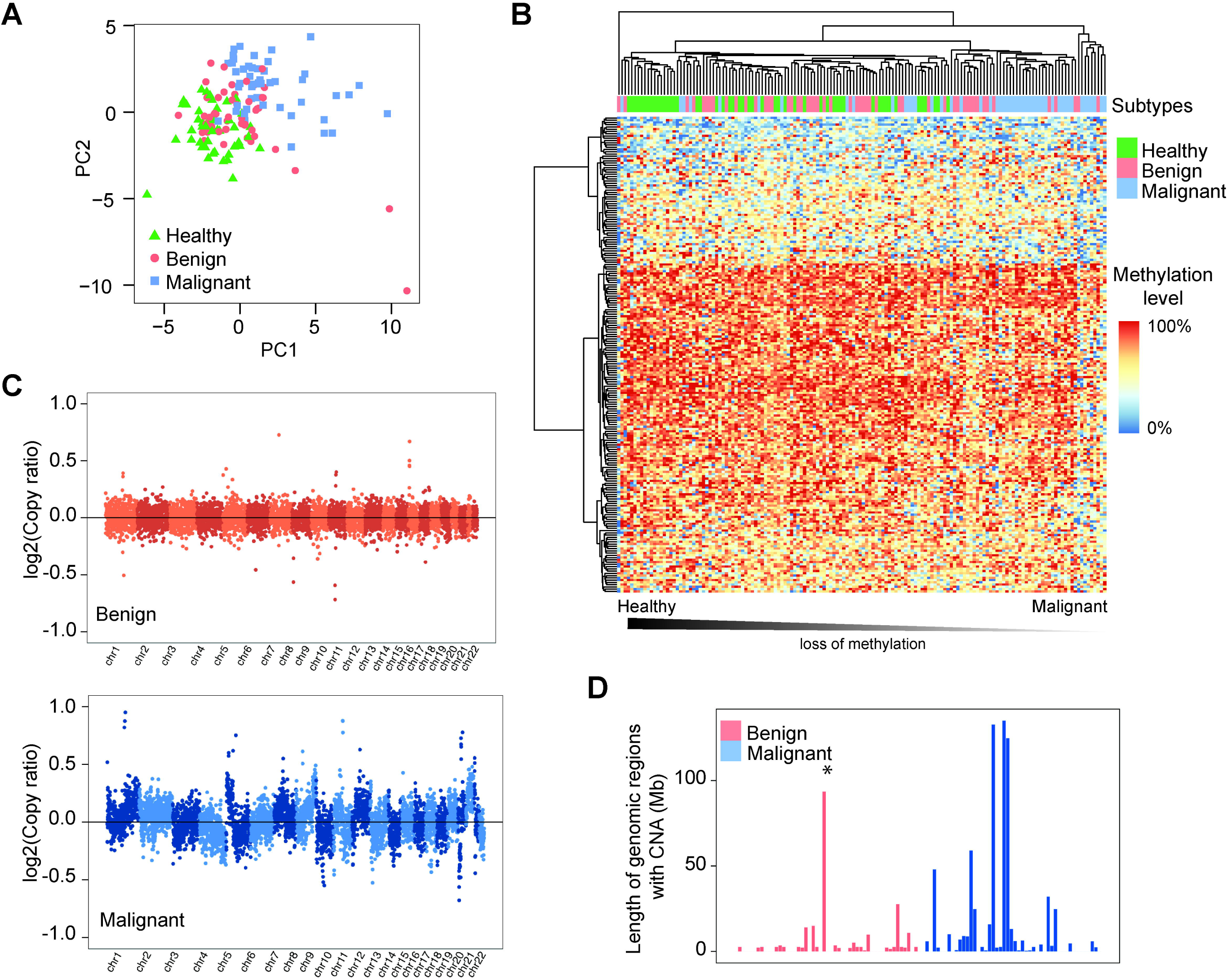
LABS-seq analysis of 5mC reveals copy number alterations in plasma samples from patients with malignant disease. A) Principal component analysis of differentially methylated regions (DMRs) in plasma from healthy individuals (n=54) and patients with benign (n=44) or malignant (n=48) disease. B) Hierarchical clustering based on 211 DMRs. Hypermethylated regions are shown in red, while hypomethylated regions are indicated in blue. C) Global copy number alterations (CNA) identified in plasma from patients with benign and malignant diseases. D) Length of genomic regions with copy number alterations for each benign and malignant sample. *Indicates a benign sample from a patient later diagnosed with cervical intraepithelial neoplasia I (CIN I).

### 5mC changes in transposable elements are predictive of malignancy

Our study thus far demonstrated that changes in 5hmC on cfDNA can predict malignancy, and since 5hmC is an intermediate product of active 5mC demethylation, we reasoned that 5mC within TEs might provide orthogonal biomarker signatures for malignancy prediction. Our analysis revealed 532 transposable element promoters that were significantly hypomethylated and associated with malignant disease, while 187 were significantly hypermethylated and associated with benign disease (**Figure 5A**). 5mC distribution enrichment analysis by TEs revealed that the majority of hypermethylation in malignant disease was found in SINE (19.25%) and Alu (42.24%), while hypomethylation was found within LINE-2 (14.09%), ERVL (11.84%), and LINE-1 (10.52%) elements was enriched in plasma from benign cases (**Figure 5B** and **Table S5**). Principal component analysis and hierarchical clustering based on 791 differentially methylated TE promoters completely separated benign from malignant cases (**Figure 5C-D** and **Table S6**). To develop a predictive model, we applied a nested cross-validation framework combined with gradient boosting (XGBoost), using feature selection based on correlation with sample labels (37,38). The model was trained on 80% of the data, while the remaining 20% were used to evaluate the model (**Figure 5E**). The resulting model achieved an AUC of 0.97 with an accuracy of 0.939 for predicting malignancy in plasma samples based on the top 25 positive and negative correlated features (**Figure 5F** and **Table S7**).

**Figure 5:**
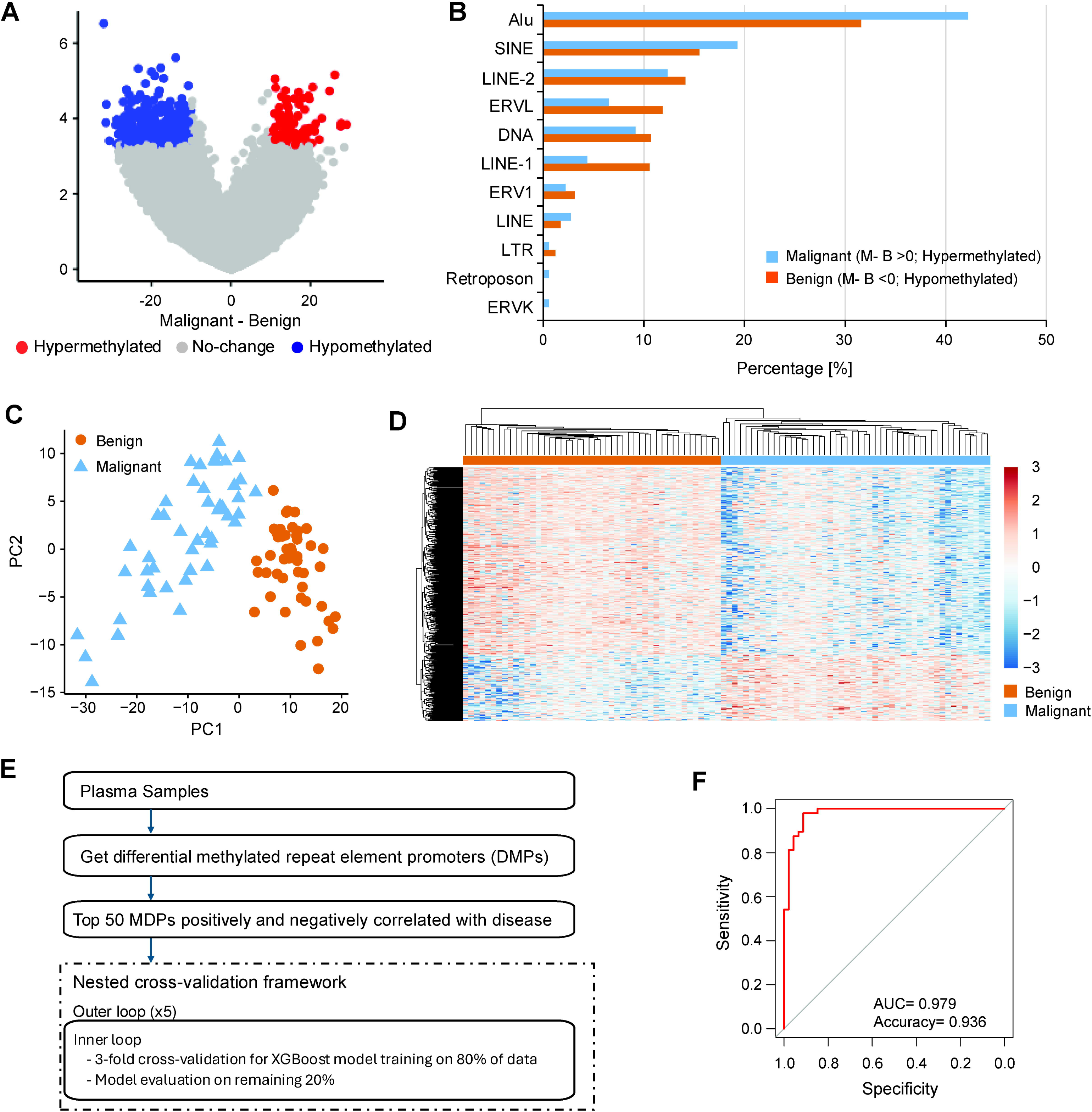
5mC signals on transposable element promoters can differentiate between benign and malignant disease. A) Volcano plot illustrating 5mC changes, characterized as hyper- and hypomethylation in the promoter regions of transposable elements between benign (n=44) and malignant (n=48) disease. In malignant disease, 532 promoters were significantly hypomethylated, while 187 promoters were significantly hypermethylated in benign disease (Student’s t-test, p < 0.0005). B) 5mC distribution enrichment by transposable elements. C) Principal component analysis based on 791 differentially methylated transposable element promoters from samples with benign and malignant disease. D) Hierarchical clustering based on differentially methylated transposable element promoters. E) Nested cross-validation framework illustrating steps involved in building a model using these differentially methylated transposable element promoters (DMPs) to distinguish patients with benign from those with malignant disease using cfDNA. F) Receiver Operating Curve with nested cross-validation employing the top 25 positively and 25 negatively correlated differentially methylated transposable element promoters.

## Discussion

The ability to accurately distinguish between benign and malignant adnexal masses remains a major clinical challenge. While 90% of adnexal masses are benign, missing a malignancy can have serious consequences due to the high mortality rate of OvCa (2–4). Additionally, current liquid biopsy assays used to evaluate the malignancy of an adnexal mass have low specificity (8,9,48). In our study, we sought to investigate the utility of 5hmC and 5mC profiling of TEs in cfDNA as a new approach to accurately and robustly distinguish benign from malignant adnexal masses.

Our study identifies two distinct 5hmC and 5mC-based signatures in TEs that can accurately differentiate between benign and malignant adnexal masses. Using the top 38 differentially hydroxymethylated TEs, our 5hmC-based model achieved an AUC of 0.893 in an independent serum cohort, demonstrating high discriminatory power across two types of analytes (plasma and serum). In parallel, our 5mC-based model, which used the top 50 highly correlated differentially methylated TE promoter regions, achieved an AUC of 0.97. This indicates that methylation profiles of TEs in cfDNA are robust biomarkers of malignancy (23,43). This finding also aligns with and extends the work of previous studies that have linked global hypomethylation and TE activation with tumorigenesis (25,44). Unlike earlier studies that focused on TE expression (17) or sequence alterations in tumor tissue (22), we demonstrate that 5hmC and 5mC modifications within specific TE classes, such as Alu, LINE-1, and LINE-2 elements, are differentially enriched in cfDNA and can provide powerful features for OvCa prediction.

Our study highlights several key points. First, the minimal feature set required for the 5hmC model (n=30) and the 5mC model (n=50) paves the way for future targeted sequencing panels that are more affordable and more scalable. Second, the effectiveness of the 5hmC model on both serum and plasma is notable, especially since serum has traditionally been the preferred analyte for many clinical chemistry tests.

Various sequencing-based approaches, such as the RNA sequencing of TE-derived transcripts (49), whole genome- and long-read sequencing (50) as well as Analysis of RepeaT EleMents in diSease (ARTEMIS) (51), have examined the role of TEs in human disease and as multicancer biomarkers (52,53). ARTEMIS, for example, captures tumor-specific changes in RE abundance and structural variation across the genome. Combined with machine learning models, ARTEMIS can predict early-stage lung or liver cancer. Our approach offers complementary information by examining the epigenetic state of these elements. Additionally, 5hmC and 5mC profiling can reveal regulatory activities and provide insights into early reprogramming events and tissue-specific enhancer dynamics that may occur before mutations and genomic instability.

More recently *Michael et al., 2025* published a study introducing the Detection of Long Interspersed Nuclear Element Altered Methylation ON plasma DNA (DIAMOND) strategy. Using a reference -free method, the DIAMOND approach uses the methylation status of CpG sites in young LINE-1 elements to predict malignancy (23). Using this approach showed that DIAMOND achieved a high classification accuracy across multiple cancer types, including OvCa. While the DIAMOND method exclusively focuses on LINE-1 methylation in the form of 5mC, our study interrogates methylation and hydroxymethylation on repeat elements across diverse TE families offering a broader view of the epigenetic landscape of TEs in OvCa. Furthermore, our models are optimized for predicting malignancy in the context of an adnexal mass, where better risk stratification methods are urgently needed.

In summary, our study supports previous research indicating that tumor development involves widespread changes in methylation and TEs dysregulation. Additionally, we demonstrate that alterations in 5hmC and 5mC on cfDNA within TEs are promising biomarkers for detecting malignancy in adnexal masses. Our findings warrant further investigation of these biomarker signatures in larger, prospective cohorts to evaluate their effectiveness either alone or combined with other existing diagnostic methods for OvCa.

## Limitations

One limitation of our study is that not all benign cases were of ovarian origin/adnexal masses; instead, they included various benign gynecologic conditions, such as fibroids and endometriosis. We also included samples from patients undergoing risk-reducing surgeries or cerclages. Overall, 12.64% of our samples were not of ovarian or adnexal origin. Although we demonstrated that 5hmC within TEs is predictive of malignancy, we used plasma samples for TE discovery and model training, while model validation was performed on serum samples. Using two different analytes (serum and plasma) might introduce unknown biases. While we evaluated the similarity of borderline and healthy samples through clustering, the small number of these samples prevented us from including them in model development. Therefore, our model is only applicable to distinguish benign from malignant samples.

## Abbreviations

5hmC: 5-hydroxymethylcytosine
5mC: 5-methylcytosine
Alu: Arthrobacter luteus
cfDNA: Cell-free DNA
CpG-Island: Palindromic cytosine-phosphate-linked-guanine islands
DhMGs: Differentially hydroxymethylated genes
DMRs: Differentially methylated regions
ERV: Endogenous retroviruses
ERVK: Endogenous retrovirus type K
ERVL: Endogenous retrovirus-like element
LABS-seq: Linear amplification-based bisulfite sequencing
LINE: Long interspersed nuclear element
LTR: Long terminal repeat
rRNA: Ribosomal RNA
SINE: Short interspersed nuclear element
snRNA: Small nuclear RNA
TE: Transposable elements
TSS: Transcription start site
TTS: Transcription termination site
UTR: Untranslated region

## Acknowledgements

The 5hmC work presented herein was supported by the University of Chicago Comprehensive Cancer Center (E.L. and C.H.). Work involving the analysis of 5mC was supported by the honorable Tina’s Wish Foundation (C.H., E.L., and M.W.). The authors would like to thank Dr. Kolla Kristjansdottir at Midwestern University for providing laboratory space to perform the LABS-seq library preparations and Dr. Pieter Faber, PhD and the staff at the University of Chicago Genomics Facility. We would also like to thank Dahlya Manning, MPH, Rohin Dhir, MD, and Calla O’Connor, MPH, for consenting patients, Leonhard Donle, PhD, for bioinformatics support, as well as all the patients who participated in this study.

## Author contributions

Conceptualization: M.W., C.H., and E.L. Data curation: M.W., X.C., and P.Z. Formal analysis: M.W., X.C., and P.Z. Funding acquisition: M.W., C.H., and E.L. Investigation: M.W., X.C., D.W.S., K.K, and S.R. Methodology: M.W., X.C., P.Z., D.W.S., K.K, and W.Z. Project administration: M.W. Resources: X.C., W.Z., C.H., and E.L. Software: X.C., P.Z., and W.Z. Supervision: W.Z., C.H., and E.L. Validation: M.W., and X.C. Visualization: M.W., X.C., and P.Z. Writing- original draft: M.W., and E.L. Writing-review and editing: M.W., X.C., P.Z., D.W.-S., K.K, S.R., W.Z., C.H., and E.L.

## Supplementary Figure Legends

**Figure S1:**
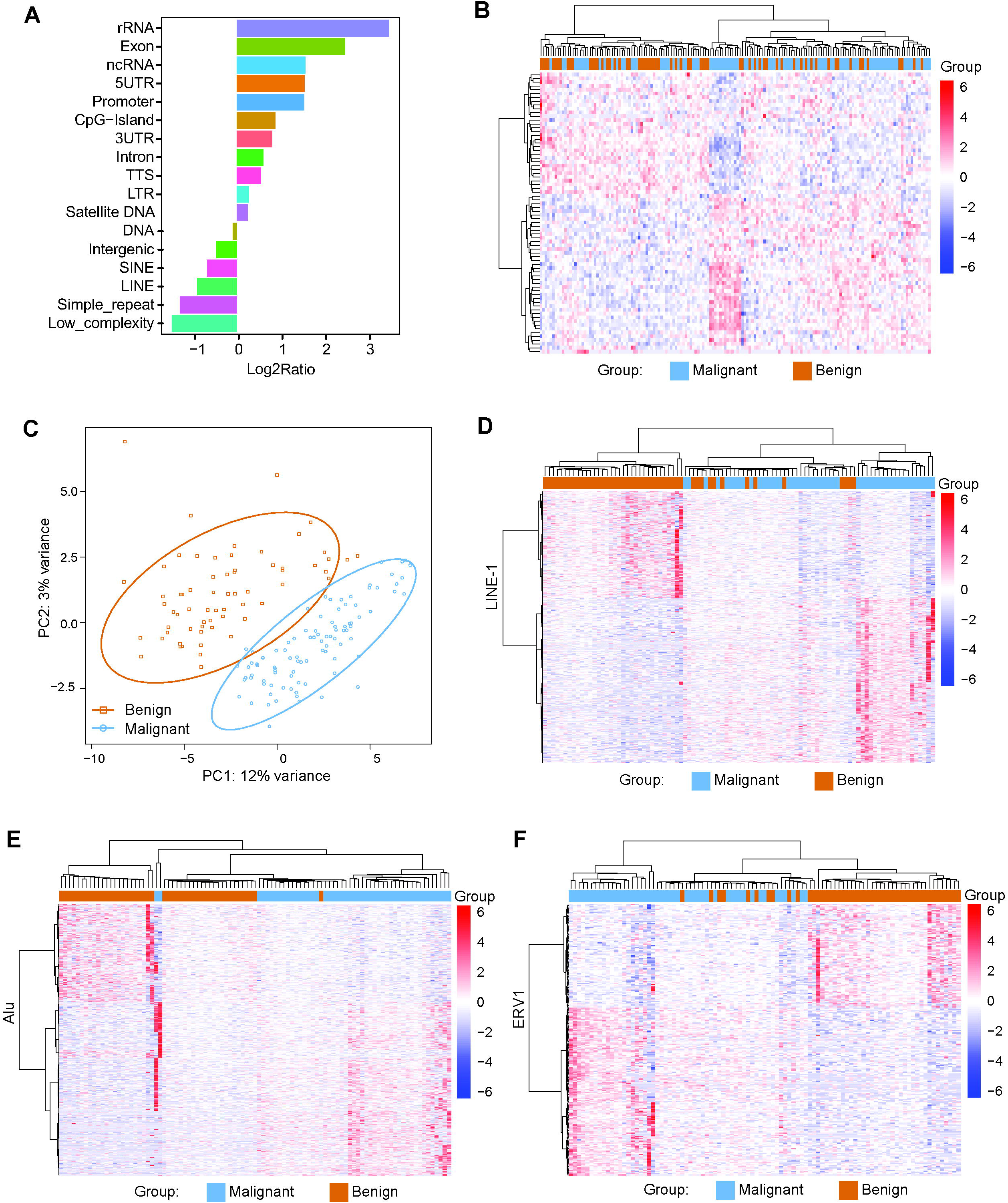
5hmC signals in plasma-derived cfDNA can differentiate between benign and malignant diseases. A) 5hmC distribution enrichment analysis by genomic features for all samples. B) Hierarchical clustering of benign and malignant samples based on 842 differentially hydroxymethylated genes (q < 0.05). C) Principal component analysis (PCA) plot of globally differentially hydroxymethylated transposable elements in benign and malignant diseases. D-F) Hierarchical clustering of benign and malignant samples based on differentially hydroxymethylated LINE-1 (379), Alu (562), and ERV1 (71) elements (q < 0.05).

**Figure S2:**
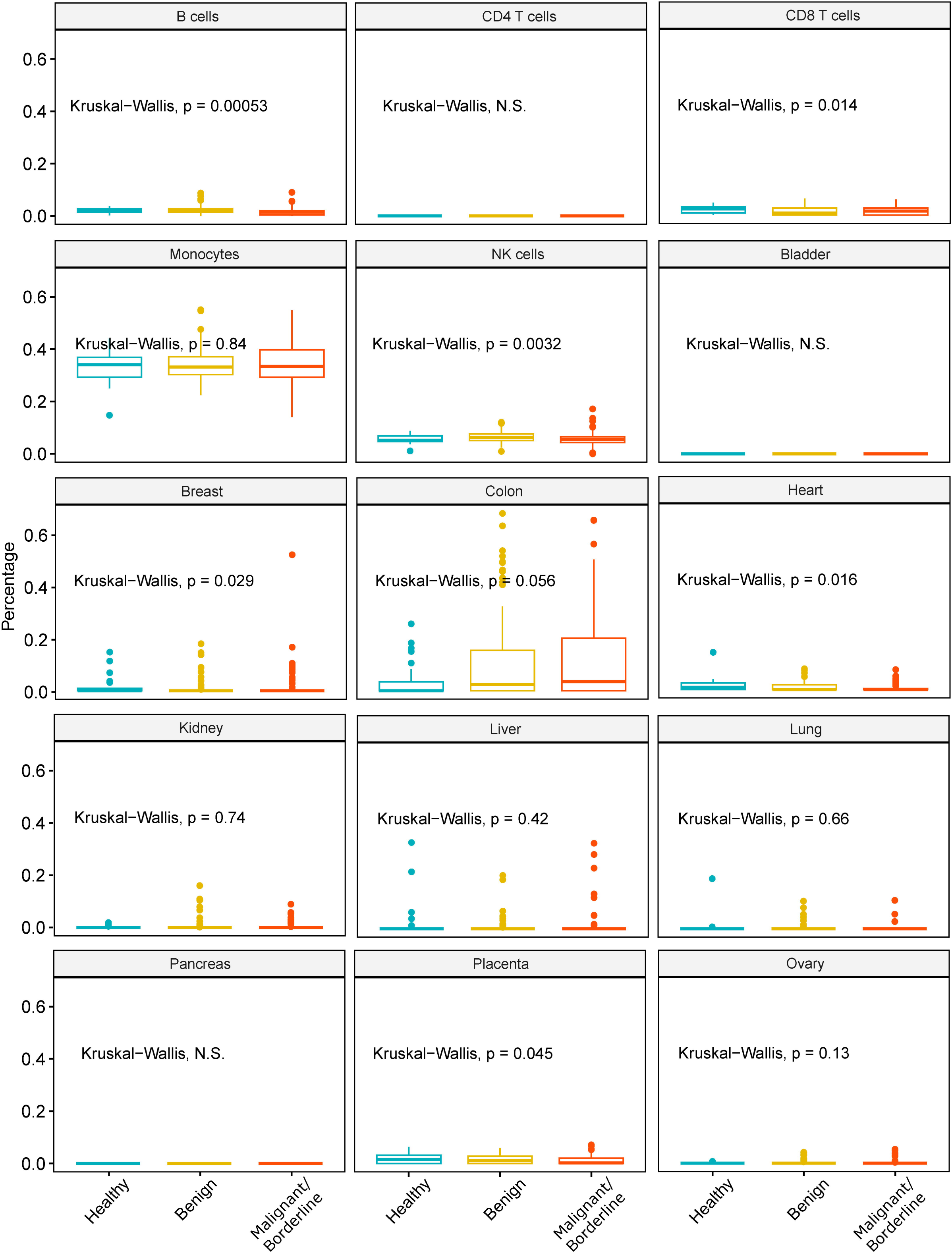
Cell deconvolution analysis of 5hmC signals in plasma-derived cfDNA. Cell deconvolution analysis based on plasma-derived 5hmC signals in cfDNA for B cells, CD4 T cells, CD8 T cells, monocytes, NK cells, bladder, breast, colon, heart, kidney, liver, lung, pancreas, placenta and ovary by healthy, benign and malignant/ borderline cases.

**Figure S3:**
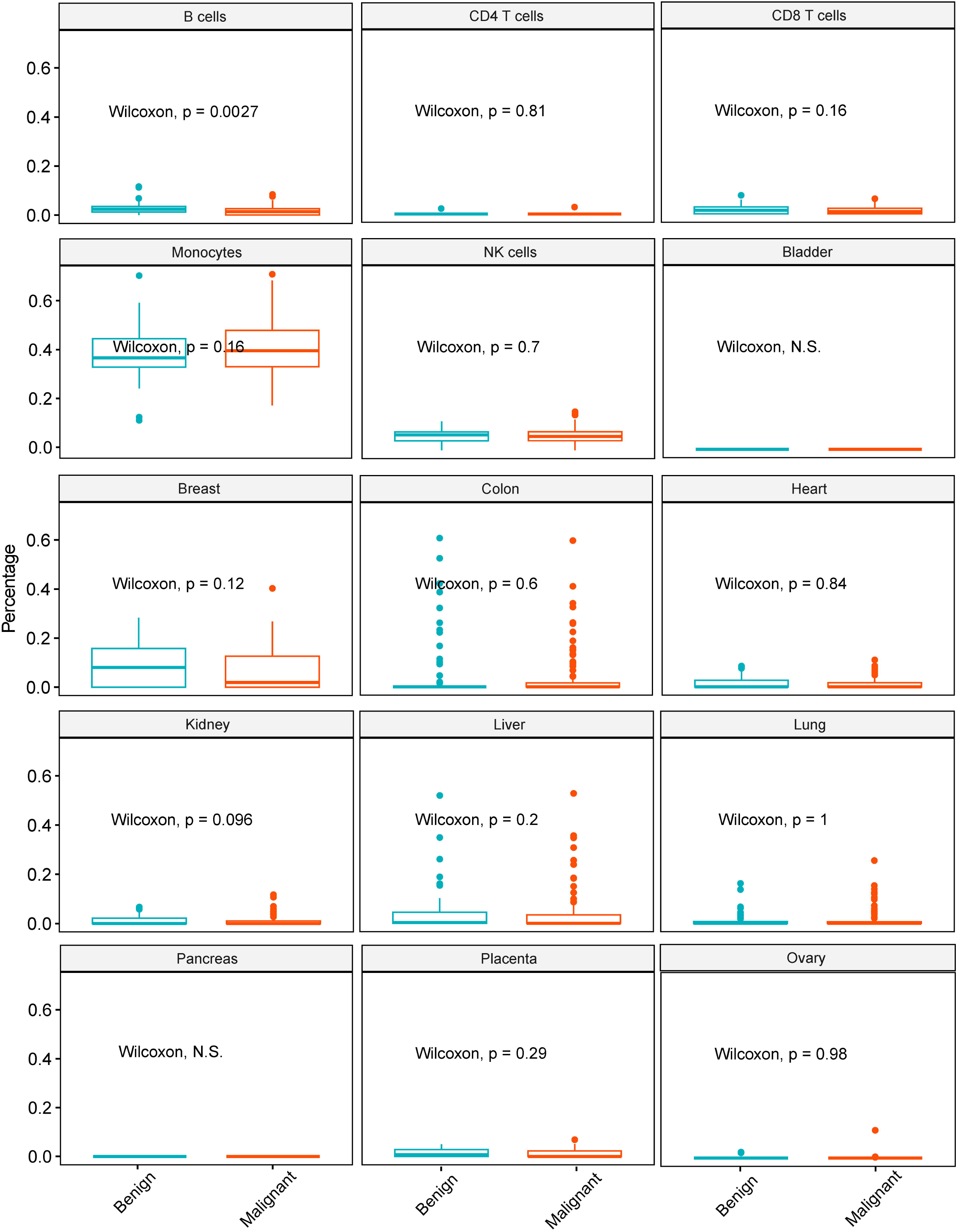
Cell deconvolution analysis of 5hmC signals in serum-derived cfDNA. Cell deconvolution analysis based on serum-derived 5hmC signals in cfDNA for B cells, CD4 T cells, CD8 T cells, monocytes, NK cells, bladder, breast, colon, heart, kidney, liver, lung, pancreas, placenta and ovary by benign and malignant cases.

**Figure S4:**
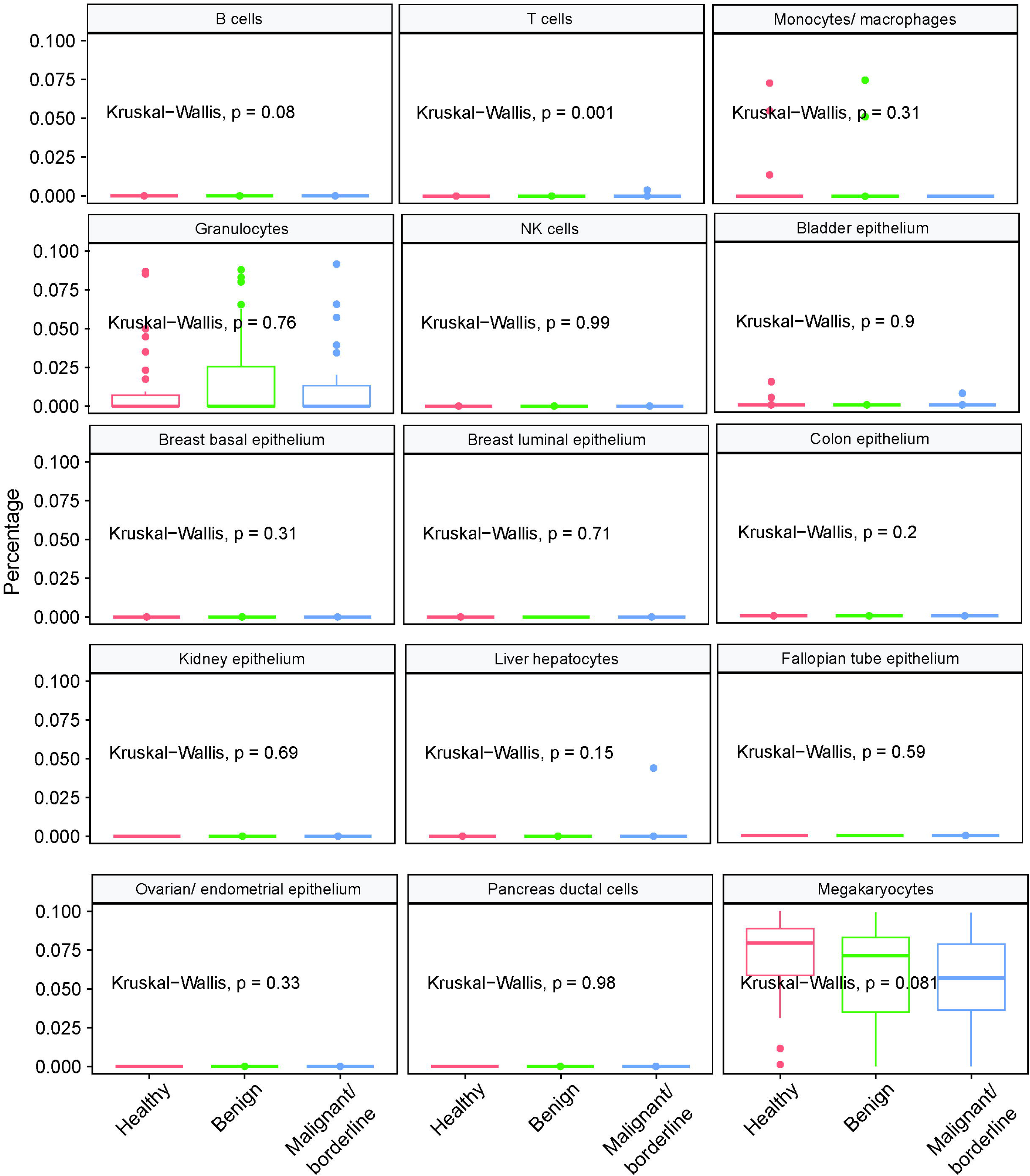
Cell deconvolution analysis of 5mC signals in plasma-derived cfDNA. Cell deconvolution analysis based on plasma-derived 5hmC signals in cfDNA for B cells, T cells, monocytes/ macrophages, NK cells, bladder epithelium, breast basal epithelium, breast luminal epithelium, colon epithelium, kidney epithelium, liver hepatocytes, fallopian tube epithelium, ovarian/ endometrial epithelium, pancreas ductal cells and megakaryocytes by healthy, benign and malignant/ borderline cases.

## Tables

Table S1: Clinicopathologic cohort overview.

Cohort tables containing clinicopathologic information for serum and plasma samples used in this study (Figure 1).

Tab1: 5hmC plasma cohort.

Tab2: 5hmC serum cohort.

Tab3: 5mC plasma cohort.

Table S2: Plasma peak distribution.

Distribution of sequencing reads by genomic features in percentage (Figure 2A).

Table S3: 5hmC distribution enrichment by repeat elements.

Enrichment of sequencing reads (%) by repeat elements (Figure 2D).

Table S4: Transposable elements used for modeling.

List of transposable elements used for building the 5hmC based model (Figure 3A).

Table S5: 5mC distribution enrichment by transposable elements.

Distribution of sequencing reads (%) by different transposable elements in benign or malignant samples (Figure 5B).

Table S6: Differentially methylated repeat element promoters.

Table containing the 339 differentially methylated (5mC) repeat element promoters, their promoter location, difference in methylation, P-values, correlation with labels, and rank of correlation (Figure 5C-D).

Table S7: Top 25 positive and negative correlated features.

Table containing the top 25 positive and negative correlated features used for 5mC-based modelling (Figure 5F).

## References

1. Sisodia RC, Del Carmen MG. Lesions of the Ovary and Fallopian Tube. N Engl J Med. 2022 Aug 25;387(8):727–36.

2. Siegel RL, Giaquinto AN, Jemal A. Cancer statistics, 2024. CA Cancer J Clin. 2024;74(1):12–49.

3. Glanc P, Benacerraf B, Bourne T, Brown D, Coleman BG, Crum C, et al. First International Consensus Report on Adnexal Masses: Management Recommendations. J Ultrasound Med Off J Am Inst Ultrasound Med. 2017 May;36(5):849–63.

4. Buys SS, Partridge E, Black A, Johnson CC, Lamerato L, Isaacs C, et al. Effect of screening on ovarian cancer mortality: the Prostate, Lung, Colorectal and Ovarian (PLCO) Cancer Screening Randomized Controlled Trial. JAMA. 2011 June 8;305(22):2295–303.

5. Carreras-Dieguez N, Glickman A, Munmany M, Casanovas G, Agustí N, Díaz-Feijoo B, et al. Comparison of HE4, CA125, ROMA and CPH-I for Preoperative Assessment of Adnexal Tumors. Diagn Basel Switz. 2022 Jan 17;12(1):226.

6. Zhang R, Siu MKY, Ngan HYS, Chan KKL. Molecular Biomarkers for the Early Detection of Ovarian Cancer. Int J Mol Sci. 2022 Oct 10;23(19):12041.

7. Cao Y, Bai Y, Yuan T, Song L, Fan Y, Ren L, et al. Single-cell bisulfite-free 5mC and 5hmC sequencing with high sensitivity and scalability. Proc Natl Acad Sci U S A. 2023 Dec 5;120(49):e2310367120.

8. Moore RG, McMeekin DS, Brown AK, DiSilvestro P, Miller MC, Allard WJ, et al. A novel multiple marker bioassay utilizing HE4 and CA125 for the prediction of ovarian cancer in patients with a pelvic mass. Gynecol Oncol. 2009 Jan;112(1):40–6.

9. Coleman RL, Herzog TJ, Chan DW, Munroe DG, Pappas TC, Smith A, et al. Validation of a second-generation multivariate index assay for malignancy risk of adnexal masses. Am J Obstet Gynecol. 2016 July;215(1):82.e1–82.e11.

10. Bristow RE, Smith A, Zhang Z, Chan DW, Crutcher G, Fung ET, et al. Ovarian malignancy risk stratification of the adnexal mass using a multivariate index assay. Gynecol Oncol. 2013 Feb;128(2):252–9.

11. Ueland FR, Desimone CP, Seamon LG, Miller RA, Goodrich S, Podzielinski I, et al. Effectiveness of a multivariate index assay in the preoperative assessment of ovarian tumors. Obstet Gynecol. 2011 June;117(6):1289–97.

12. Jacobs I, Oram D, Fairbanks J, Turner J, Frost C, Grudzinskas JG. A risk of malignancy index incorporating CA 125, ultrasound and menopausal status for the accurate preoperative diagnosis of ovarian cancer. Br J Obstet Gynaecol. 1990 Oct;97(10):922–9.

13. Urban RR, Pappas TC, Bullock RG, Munroe DG, Bonato V, Agnew K, et al. Combined symptom index and second-generation multivariate biomarker test for prediction of ovarian cancer in patients with an adnexal mass. Gynecol Oncol. 2018 Aug;150(2):318–23.

14. Fritsche HA, Bullock RG. A reflex testing protocol using two multivariate index assays improves the risk assessment for ovarian cancer in patients with an adnexal mass. Int J Gynaecol Obstet Off Organ Int Fed Gynaecol Obstet. 2023 Aug;162(2):485–92.

15. Nishiyama A, Nakanishi M. Navigating the DNA methylation landscape of cancer. Trends Genet TIG. 2021 Nov;37(11):1012–27.

16. Vasanthakumar A, Godley LA. 5-hydroxymethylcytosine in cancer: significance in diagnosis and therapy. Cancer Genet. 2015 May;208(5):167–77.

17. Pisanic TR II, Wang Y, Sun H, Considine M, Li L, Wang TH, et al. Methylomic Landscapes of Ovarian Cancer Precursor Lesions. Clin Cancer Res. 2020 Dec 1;26(23):6310–20.

18. Chennakesavalu M, Moore K, Chaves G, Veeravalli S, TerHaar R, Wu T, et al. 5-Hydroxymethylcytosine Profiling of Cell-Free DNA Identifies Bivalent Genes That Are Prognostic of Survival in High-Risk Neuroblastoma. JCO Precis Oncol. 2024 Jan;8:e2300297.

19. Chiu BCH, Chen C, You Q, Chiu R, Venkataraman G, Zeng C, et al. Alterations of 5-hydroxymethylation in circulating cell-free DNA reflect molecular distinctions of subtypes of non-Hodgkin lymphoma. NPJ Genomic Med. 2021 Feb 11;6(1):11.

20. Cui XL, Nie J, Zhu H, Kowitwanich K, Beadell AV, West-Szymanski DC, et al. LABS: linear amplification-based bisulfite sequencing for ultrasensitive cancer detection from cell-free DNA. Genome Biol. 2024 June 14;25(1):157.

21. Weigert M, Cui XL, West-Szymanski D, Yu X, Bilecz AJ, Zhang Z, et al. 5-Hydroxymethylcytosine signals in serum are a predictor of chemoresistance in high-grade serous ovarian cancer. Gynecol Oncol. 2024 Mar;182:82–90.

22. Rodriguez-Martin B, Alvarez EG, Baez-Ortega A, Zamora J, Supek F, Demeulemeester J, et al. Pan-cancer analysis of whole genomes identifies driver rearrangements promoted by LINE-1 retrotransposition. Nat Genet. 2020 Mar;52(3):306–19.

23. Michel M, Heidary M, Mechri A, Da Silva K, Gorse M, Dixon V, et al. Noninvasive Multicancer Detection Using DNA Hypomethylation of LINE-1 Retrotransposons. Clin Cancer Res. 2025 Apr 1;31(7):1275–91.

24. C MD, Kh B. LINE-1 retrotransposition and its deregulation in cancers: implications for therapeutic opportunities. Genes Dev [Internet]. 2023 Dec 26 [cited 2025 June 5];37(21–24). Available from: https://pubmed.ncbi.nlm.nih.gov/38092519/

25. Burns KH. Transposable elements in cancer. Nat Rev Cancer. 2017 July;17(7):415–24.

26. Song CX, Yin S, Ma L, Wheeler A, Chen Y, Zhang Y, et al. 5-Hydroxymethylcytosine signatures in cell-free DNA provide information about tumor types and stages. Cell Res. 2017 Oct;27(10):1231–42.

27. Babraham Bioinformatics - FastQC A Quality Control tool for High Throughput Sequence Data [Internet]. [cited 2025 June 30]. Available from: https://www.bioinformatics.babraham.ac.uk/projects/fastqc/

28. Babraham Bioinformatics - Trim Galore! [Internet]. [cited 2025 June 30]. Available from: https://www.bioinformatics.babraham.ac.uk/projects/trim_galore/

29. Li H, Handsaker B, Wysoker A, Fennell T, Ruan J, Homer N, et al. The Sequence Alignment/Map format and SAMtools. Bioinforma Oxf Engl. 2009 Aug 15;25(16):2078–9.

30. Fishilevich S, Nudel R, Rappaport N, Hadar R, Plaschkes I, Iny Stein T, et al. GeneHancer: genome-wide integration of enhancers and target genes in GeneCards. Database J Biol Databases Curation. 2017 Jan 1;2017:bax028.

31. Karolchik D, Hinrichs AS, Furey TS, Roskin KM, Sugnet CW, Haussler D, et al. The UCSC Table Browser data retrieval tool. Nucleic Acids Res. 2004 Jan 1;32(Database issue):D493–496.

32. Liao Y, Smyth GK, Shi W. featureCounts: an efficient general purpose program for assigning sequence reads to genomic features. Bioinforma Oxf Engl. 2014 Apr 1;30(7):923–30.

33. Varet H, Brillet-Guéguen L, Coppée JY, Dillies MA. SARTools: A DESeq2- and EdgeR-Based R Pipeline for Comprehensive Differential Analysis of RNA-Seq Data. PloS One. 2016;11(6):e0157022.

34. Zhou Y, Zhou B, Pache L, Chang M, Khodabakhshi AH, Tanaseichuk O, et al. Metascape provides a biologist-oriented resource for the analysis of systems-level datasets. Nat Commun. 2019 Apr 3;10(1):1523.

35. Krueger F, Andrews SR. Bismark: a flexible aligner and methylation caller for Bisulfite-Seq applications. Bioinforma Oxf Engl. 2011 June 1;27(11):1571–2.

36. Heinz S, Benner C, Spann N, Bertolino E, Lin YC, Laslo P, et al. Simple combinations of lineage-determining transcription factors prime cis-regulatory elements required for macrophage and B cell identities. Mol Cell. 2010 May 28;38(4):576–89.

37. Chen T, Guestrin C. XGBoost: A Scalable Tree Boosting System. In: Proceedings of the 22nd ACM SIGKDD International Conference on Knowledge Discovery and Data Mining [Internet]. New York, NY, USA: Association for Computing Machinery; 2016 [cited 2025 July 25]. p. 785–94. (KDD’16). Available from: https://dl.acm.org/doi/10.1145/2939672.2939785

38. Parvandeh S, Yeh HW, Paulus MP, McKinney BA. Consensus features nested cross-validation. Bioinformatics. 2020 Jan 27;36(10):3093–8.

39. Nakauchi Y, Azizi A, Thomas D, Corces MR, Reinisch A, Sharma R, et al. The Cell Type–Specific 5hmC Landscape and Dynamics of Healthy Human Hematopoiesis and TET2-Mutant Preleukemia. Blood Cancer Discov. 2022 July 6;3(4):346–67.

40. Cui XL, Nie J, Ku J, Dougherty U, West-Szymanski DC, Collin F, et al. A human tissue map of 5-hydroxymethylcytosines exhibits tissue specificity through gene and enhancer modulation. Nat Commun. 2020 Dec 2;11(1):6161.

41. Loyfer N, Magenheim J, Peretz A, Cann G, Bredno J, Klochendler A, et al. A DNA methylation atlas of normal human cell types. Nature. 2023 Jan;613(7943):355–64.

42. Applebaum MA, Barr EK, Karpus J, Nie J, Zhang Z, Armstrong AE, et al. 5-Hydroxymethylcytosine Profiles Are Prognostic of Outcome in Neuroblastoma and Reveal Transcriptional Networks That Correlate With Tumor Phenotype. JCO Precis Oncol. 2019;3:PO.18.00402.

43. Kanholm T, Rentia U, Hadley M, Karlow JA, Cox OL, Diab N, et al. Oncogenic Transformation Drives DNA Methylation Loss and Transcriptional Activation at Transposable Element Loci. Cancer Res. 2023 Aug 1;83(15):2584–99.

44. Rodic N. LINE-1 activity and regulation in cancer. Front Biosci Landmark Ed. 2018 Mar 1;23(9):1680–6.

46. Dawson MA, Kouzarides T. Cancer epigenetics: from mechanism to therapy. Cell. 2012 July 6;150(1):12–27.

47. Bell D, Berchuck A, Birrer M, Chien J, Cramer DW, Dao F, et al. Integrated genomic analyses of ovarian carcinoma. Nature. 2011 June;474(7353):609–15.

48. Van Calster B, Timmerman D, Bourne T, Testa AC, Van Holsbeke C, Domali E, et al. Discrimination Between Benign and Malignant Adnexal Masses by Specialist Ultrasound Examination Versus Serum CA-125. JNCI J Natl Cancer Inst. 2007 Nov 21;99(22):1706–14.

49. McKerrow W, Wang X, Mendez-Dorantes C, Mita P, Cao S, Grivainis M, et al. LINE-1 expression in cancer correlates with p53 mutation, copy number alteration, and S phase checkpoint. Proc Natl Acad Sci U S A. 2022 Feb 22;119(8):e2115999119.

50. Rybacki K, Xia M, Ahsan MU, Xing J, Wang K. Assessing the Expression of Long Interspersed Elements (LINEs) via Long-Read Sequencing in Diverse Human Tissues and Cell Lines. Genes. 2023 Sept 29;14(10):1893.

51. Annapragada AV, Niknafs N, White JR, Bruhm DC, Cherry C, Medina JE, et al. Genome-wide repeat landscapes in cancer and cell-free DNA. Sci Transl Med. 2024 Mar 13;16(738):eadj9283.

52. Doucet-O’Hare TT, Rodić N, Sharma R, Darbari I, Abril G, Choi JA, et al. LINE-1 expression and retrotransposition in Barrett’s esophagus and esophageal carcinoma. Proc Natl Acad Sci. 2015 Sept;112(35):E4894–900.

53. Steranka JP, Tang Z, Grivainis M, Huang CRL, Payer LM, Rego FOR, et al. Transposon insertion profiling by sequencing (TIPseq) for mapping LINE-1 insertions in the human genome. Mob DNA. 2019 Mar 8;10(1):8.

